# Determination of the practical utility of ESMO Scale for Clinical Actionability of molecular Targets (ESCAT): mapping OncoKB level 1 alterations using ESCAT

**DOI:** 10.64898/2026.05.16.26353390

**Authors:** M Kordes, D Chakravarty, E Boberg, M Creignou, L De Petris, C Karlsson, L Liu Burström, SP Suehnholz, J Yachnin, OPB Wiklander, F Haglund de Flon

## Abstract

**Background:** The European Society for Medical Oncology (ESMO) Scale for Clinical Actionability of molecular Targets (ESCAT) ranks genomic alterations by the evidence supporting the predictive value of the molecular target for response to targeted therapies. No openly available, systematically curated set of standard care biomarkers mapped to the ESCAT framework exists to support clinical decision-making or harmonize biomarker interpretation.

**Methods:** We mapped all OncoKB™ Level 1 biomarkers to ESCAT tiers using evidence cited by OncoKB™, excluding abstract-only data. Eight board-certified oncologists and hematologists independently assigned ESCAT tiers, with discrepancies resolved through structured consensus meetings. Recurring evidence scenarios that did not correspond to any existing ESCAT tier informed a set of a priori–defined modifications, which were subsequently applied to biomarkers that could not be classified using native ESCAT criteria.

**Results:** Of 188 OncoKB™ Level 1 biomarkers, 16 were excluded due to abstract-only evidence. Using native ESCAT criteria, 51% of the remaining biomarkers were classified as Tier 1, 3% Tier 2, 18% Tier 3, 6% Tier X and 22% could not be assigned to any tier. Applying the modified ESCAT criteria resolved all previously unclassifiable biomarkers and increased Tier 1 assignments to 73%. Inter-rater reliability (Krippendorff’s α) was moderate (0.586) and 62% of classifications required consensus discussions. Comparison with ESCAT tiers reported in ESMO Clinical Practice Guidelines showed improved concordance when using the modified criteria.

**Conclusions:** The native ESCAT criteria are highly stringent, resulting in many FDA-recognized, clinically validated biomarkers that are currently assigned level 1 by OncoKB™ not mapping to any existing tier. Our predefined modifications improved alignment with OncoKB™ Level 1 designations and with published ESMO clinical practice guidelines. The mapped set of standard care biomarkers are provided on the OncoKB™ website, offering a practical resource that harmonizes ESCAT tiers of evidence with a widely adopted levels of evidence schema.

**SUMMARY:** *Key objective:* Can FDA-recognized OncoKB Level 1 biomarkers be systematically mapped to ESCAT tiers using curated evidence alone, and how can ESCAT be operationally adapted to resolve recurrent evidence scenarios that prevent classification?

*Knowledge generated:* Using native ESCAT criteria, 22% of OncoKB Level 1 biomarkers could not be assigned to any tier, and inter-rater agreement was moderate. Predefined modifications resolved all unmappable cases and increased Tier I classifications from 51% to 73%, with improved concordance to ESMO Clinical Practice Guidelines.

*Relevance:* The resulting harmonized ESCAT classification provides a structured and reproducible resource for molecular tumor boards and clinical sequencing pipelines, enabling more consistent interpretation of actionable biomarkers. The proposed modifications clarify how to handle common evidence scenarios, improving alignment between regulatory, guideline, and evidence-based frameworks in precision oncology.

**Highlights:**

- OncoKB Level 1 biomarkers was mapped to ESCAT tiers using OncoKB evidence as of 29 Sept 2025.
- Using native ESCAT criteria, 22% of validated Level 1 biomarkers could not be assigned to any tier.
- Modified ESCAT criteria resolved unmappable biomarkers and increased Tier 1 assignments from 51% to 73%.

## INTRODUCTION

Standardized frameworks for evaluating the clinical relevance of genomic alterations are critical to guide therapeutic decision-making. Among the most widely used systems are the European Society for Medical Oncology Scale for Clinical Actionability of molecular Targets (ESCAT) and the OncoKB™ Therapeutic Levels of Evidence framework [1,2]. Both define the strength of evidence linking genomic alterations to drug response, yet they differ in scope, structure, and clinical adoption.

ESCAT ranks genomic alterations into tiers based solely on the quality and rigor of evidence demonstrating that an alteration predicts benefit from a matched targeted therapy, ranging from Tier I (ready for routine clinical use) to Tier X (no evidence of actionability). Sublevels (e.g., I-A, I-B, I-C) further refine these tiers by incorporating regulatory approval status, clinical trial design features, and magnitude of clinical benefit [3]. In contrast, the OncoKB™ framework provides a partially regulatory-anchored hierarchy of actionability, particularly for standard care biomarkers (OncoKB™ Levels 1 and 2). Level 1 biomarkers are defined by whether the US Food and Drug Administration (FDA) drug label for a precision oncology drug requires the biomarker as a patient-selection criterion for treatment. Biomarkers are assigned OncoKB™ level 2 if included in indication-specific professional guidelines, such as the National Cancer Compendium Network (NCCN), as recommended criteria for patient selection for a targeted therapy. Investigational biomarkers are assigned based on the type and strength of cancer type–specific clinical (Level 3A) or laboratory (Level 4) evidence supporting the biomarker as predictive of response to a targeted therapy and outside the scope of this study.

Despite conceptual overlap, the two systems are not directly interoperable. OncoKB™ incorporates regulatory status and guideline inclusion into its standard care designations, whereas ESCAT applies methodological criteria across all tiers, prioritizing prospective trial design and clinical benefit. As a result, many biomarkers considered Level 1 standard care in OncoKB™ do not map to any ESCAT tier, particularly when the FDA drug labels are supported by single-arm studies, aggregated biomarker groups, or trials lacking predefined ESCAT endpoints. Furthermore, ESCAT classifications remain incomplete across cancer types in published ESMO Clinical Practice Guidelines, and no openly available resource exists that systematically curates ESCAT tiers for clinical or research use.

To address this gap, we systematically mapped all OncoKB™ Level 1 biomarkers as of 29 September 2025 to ESCAT tiers using only the rigorously curated evidence cited within OncoKB™. We identified recurrent evidence patterns that did not align with existing ESCAT criteria and prospectively defined a set of modifications to improve applicability of ESCAT (which we refer to as “native” ESCAT). We present the resulting harmonized (termed “modified”) ESCAT-aligned dataset, outline structural constraints of the current (native) ESCAT framework, and propose pragmatic refinements to support more consistent and clinically aligned use of ESCAT in precision oncology.

## METHODS

### Evidence Source and Classification Principles

To assign ESCAT Tiers to OncoKB™ level 1 molecular targets, we exclusively relied upon evidence curated within the OncoKB™ knowledge base. Although some curated entries referenced meeting abstracts, for the purposes of this study, only peer-reviewed evidence that could be critically appraised in full was included and evidence available solely as abstracts was excluded. No literature beyond the OncoKB-referenced evidence set was used. The evidence dataset was frozen at the start of the first curation round (May 2024) and updated and re-evaluated in a second review (September 29, 2025).

Several referenced trials evaluated composite biomarkers comprising multiple genes linked by shared biological functions (e.g., homologous recombination deficiency [HRD]-associated genes). In such cases, OncoKB™ listed FDA-approved biomarkers despite limited patient-level data for some constituent genes. Given that clinical benefit in composite biomarker cohorts was likely driven by a subset of high-impact genes (e.g., *BRCA2* in the HRD context), ESCAT Tier I evidence was assigned only to the genes with clear clinical actionability. Remaining genes were classified as ESCAT Tier III-B in the absence of subgroup-specific analyses.

For biomarkers supported by prospective non-randomized studies, clinical benefit was evaluated using the ESMO Magnitude of Clinical Benefit Scale (ESMO-MCBS for solid tumors; ESMO-MCBS-H for hematologic malignancies) [3]. When an official MCBS scorecard was available, it was applied directly. For 12 biomarkers without previously published scorecards, one reviewer completed the corresponding MCBS/MCBS-H form during consensus review (Supplementary Table S1). Several studies reporting survival endpoints did not meet MCBS thresholds for clinical benefit due to missing quality-of-life data. These studies did not fulfill criteria for ESCAT Tier I-B, II-B nor did they map to any other ESCAT tier.

To provide users with a living document of the current OncoKB level 1 to ESCAT mappings, the dataset of mapped genomic alterations and ESCAT-tier assignments generated in this study will be made publicly available in the “Actionable Genes” page of the oncokb.org website in conjunction with publication of this manuscript.

### Reviewer Panel and Training

Eight board-certified oncologists and hematologists from the Precision Cancer Medicine program at Karolinska Institute and the Molecular Tumor Boards at Karolinska University Hospital served as reviewers. All reviewers completed standardized training in ESCAT criteria and the relevant OncoKB™ standard operating procedures before scoring.

### Pilot Calibration of ESCAT Scoring

A pilot run of six OncoKB™ Level 1 biomarkers was conducted to calibrate scoring. All eight reviewers independently assigned ESCAT tiers. Agreement required identical tier and sublevel assignment (e.g., I-B ≠ I-C). Complete agreement was achieved in one case, near agreement (7 of 8 reviewers) in two cases, and partial agreement (<7 of 8 reviewers) in three cases. All discordant or unmapped biomarkers were discussed to reconcile interpretative differences, and a standard operating protocol was established. Pilot scores were excluded from the final dataset, and all biomarkers were re-evaluated during the main scoring phase. Given the variability in the pilot phase, each biomarker in the main scoring phase was evaluated by three independent reviewers.

### Structured Scoring and Consensus Protocol

Biomarkers were reviewed in random order and randomly assigned to three reviewers, who independently submitted ratings to the non-scoring coordinator (FHdF). The coordinator compiled ratings in a central database, accessible to all reviewers once all three scores for a biomarker had been submitted.

If all three reviewers assigned the same ESCAT tier and sublevel, the classification was finalized. Any disagreement or inability to match a biomarker to a tier triggered discussion at bi-weekly consensus meetings with a quorum of ≥4 reviewers. Any reviewer could escalate a biomarker for discussion regardless of concordance if uncertainty existed regarding ESCAT interpretation. Consensus meetings continued until full agreement was reached for all contested classifications.

Biomarkers that did not map to any ESCAT tier under native criteria were subsequently re-evaluated using predefined modified ESCAT criteria (Figure 1A). During pilot calibration, the reviewer groups identified recurrent evidence patterns that did not map to any native ESCAT tier. Before beginning the main scoring phase, the reviewer group defined a set of a priori, clinically aligned modifications to the ESCAT criteria to enable consistent classification of such cases.

**Figure 1.**
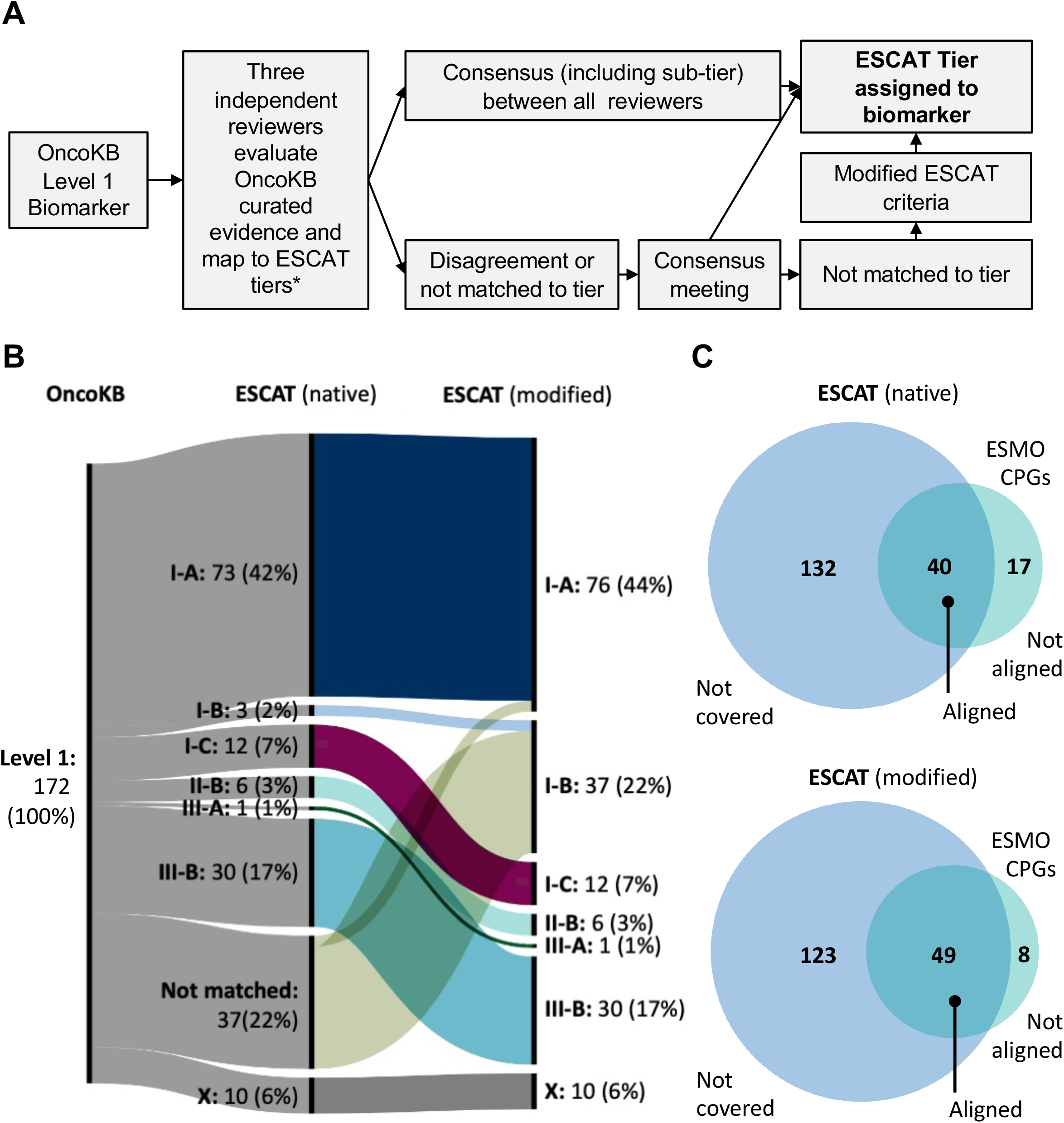
Workflow, classification outcomes, and concordance of native and modified ESCAT criteria. (A) Schematic overview of the ESCAT mapping workflow. Each OncoKB™ Level 1 biomarker was independently evaluated by three reviewers using only the evidence curated within OncoKB. Biomarkers supported exclusively by abstract-only evidence were excluded from further analysis (*). Concordant classifications were accepted directly. Biomarkers with discrepant ratings or without a compatible native ESCAT tier were discussed at consensus meetings. Biomarkers remaining unmatched under native criteria were re-evaluated using predefined modified ESCAT criteria, resulting in a final tier assignment for all biomarkers. (B) Sankey diagram illustrating the distribution of OncoKB™ Level 1 biomarkers across native and modified ESCAT tiers. Under native ESCAT criteria, 22% of biomarkers did not match any tier. Applying the predefined modifications resolved all non-matched biomarkers and increased Tier I assignments from 51% to 73%, primarily through reclassification into Tier I-A and I-B. (C) Concordance between ESCAT tier assignments and ESMO Clinical Practice Guidelines (CPGs). Venn diagrams show overlap between ESCAT classifications derived using native (top) and modified (bottom) criteria and ESCAT tiers explicitly assigned in CPGs. Of 57 OncoKB™ Level 1 biomarkers represented in CPGs, concordance increased from 40 (70%) under native ESCAT to 49 (86%) under modified ESCAT. The number of non-aligned assignments decreased from 17 to 8 following application of the modified criteria.

During the pilot calibration phase, the reviewer group identified recurrent evidence scenarios in which the published ESCAT criteria did not provide a compatible tier assignment despite the presence of clinically established biomarker–drug associations. Before initiation of the main scoring phase, the reviewer panel therefore defined two predefined operational modifications to enable consistent classification of such cases (The specific scenarios and their rationale are described in full in the Results).

First, single-arm studies reporting survival outcomes but lacking the quality-of-life data required for formal ESMO-MCBS threshold determination were adjudicated for substantial clinical benefit through structured expert consensus, rather than being left unclassifiable under native ESCAT criteria.

Second, biomarker–drug pairs supported by studies demonstrating outcomes comparable, but not superior, to an already established alteration-matched standard-of-care therapy were allowed to map to ESCAT Tier I-A within the operational framework used in this study.

These operational rules were defined during pilot calibration and applied prospectively during the main scoring phase only to biomarkers that could not be classified using native ESCAT criteria. These operational rules were applied only to biomarkers that could not be assigned to an ESCAT tier using the native criteria. Application of these rules to biomarkers already classified under native ESCAT criteria was evaluated during internal quality-control analyses and did not result in any changes to their tier assignments.

### Systematic Review of ESMO Clinical Practice Guidelines

All ESMO Clinical Practice Guidelines (CPGs) available as of 26 August 2025 were manually reviewed, excluding supportive/palliative care, COVID-19 guidance, living documents, and consensus statements. Each CPG (including supplementary materials when available) was screened for explicit ESCAT assignments. Biomarker, matched therapy, ESCAT tier, and guideline source were tabulated.

### Statistical Analysis

Statistical analyses were conducted in RStudio (version 2025.05.0+496) using the packages *ggplot2, dplyr, readxl*, and *irr*.

## RESULTS

### Native ESCAT Classification

Of 188 OncoKB™ Level 1 biomarkers, 16 were supported exclusively by abstract data and were excluded. The remaining 172 biomarkers were evaluated using native ESCAT criteria. Under strict application of these criteria, 87 biomarkers (51%) mapped to Tier I, including 73 Tier I-A (42%), 3 Tier I-B (2%), and 12 Tier I-C (7%). Six biomarkers (3%) were classified as Tier II (all Tier II-B). A total of 31 biomarkers (18%) were assigned to Tier III, comprising one Tier III-A (1%) and 30 Tier III-B (17%). Ten biomarkers (6%) mapped to Tier X, while none fulfilled criteria for Tier IV or Tier V. In total, 37 biomarkers (22%) did not map to any ESCAT tier (Figure 1B; Supplementary Table S2).

Thus, nearly one-quarter of OncoKB™ Level 1 biomarkers, representing FDA-recognized and clinically validated standard care biomarkers, did not meet the methodological thresholds required for ESCAT tier assignment under the native framework.

### Evidence Patterns Underlying Non-Mapping Under Native ESCAT Criteria

Two recurrent evidence scenarios accounted for all 37 biomarkers (22%) that could not be classified using native ESCAT criteria. First, several biomarkers were supported by prospective single-arm studies that reported survival outcomes but lacked the quality-of-life data required to reach the ESMO-MCBS threshold for substantial clinical benefit (MCBS and MCBS-H Evaluation Form 3) required for Tier I-B; the presence of survival data also precludes Tier II-B assignment. An example is vemurafenib for BRAF V600E– mutated Erdheim–Chester disease, for which marked clinical and survival benefit has been demonstrated in single-arm studies that did not report formal quality-of-life endpoints.

Second, some therapies demonstrated survival outcomes comparable, but not superior, to established alteration-matched standard care treatments. Under native ESCAT criteria, absence of superior outcomes prevents Tier I-A classification, while the reporting of survival endpoints again precludes Tier II-B. An example is dasatinib in BCR::ABL1-positive chronic myeloid leukemia, for which overall survival was comparable to imatinib.

### Impact of Modified ESCAT Criteria on Biomarker Classification

We recognized these two evidence limitations early during reviewer training and pilot calibration and therefore defined two clinically aligned modifications to enable consistent classification of OncoKB™ Level 1 biomarkers falling into these evidence scenarios: First, for single-arm trials that reported survival outcomes but lacked quality-of-life data, we relaxed the formal ESMO-MCBS threshold and evaluated clinical benefit by structured expert consensus. This enabled assignment of 35 previously unmapped biomarkers to Tier I-B. Second, we classified biomarkers supported by studies demonstrating survival outcomes comparable to alteration-matched standard-care as Tier I-A, allowing assignment of 3 previously unmapped biomarkers to this tier.

Applying these predefined modifications resolved all 37 biomarkers (22%) that were previously unmapped under native ESCAT criteria. After reclassification, all 172 biomarkers were assigned to an ESCAT tier. Under the modified framework, 76 biomarkers (44%) were classified as Tier I-A, 37 (22%) as Tier I-B, and 12 (7%) as Tier I-C. Assignment to lower tiers were unchanged, with 6 biomarkers (3%) assigned to Tier II-B, 1 (1%) to Tier III-A, 30 (17%) to Tier III-B, and 10 (6%) to Tier X. No biomarkers met criteria for Tier IV or V, and none remained unmapped (Figure 1B; Supplementary Table S2).

In summary, these modifications increased the proportion of Tier I classifications from 51% (87/172) under native ESCAT to 73% (125/172) under the modified criteria. Despite this increase, 47 (27%) of OncoKB™ Level 1 biomarkers remained within Tiers II–X, predominantly reflecting composite biomarkers or those without demonstrated clinical benefit.

### Concordance with ESMO Clinical Practice Guidelines

To evaluate the external validity of the native and modified ESCAT classifications, we examined all OncoKB™ Level 1 biomarkers that had an explicit ESCAT tier assignment in CPGs. Of the 172 Level 1 biomarkers evaluated, 57 (33%) were represented in ESMO CPGs with an assigned ESCAT tier. We also identified 23 ESCAT-tiered biomarkers present in CPGs but not classified as OncoKB™ Level 1; these were catalogued separately and excluded from concordance analyses (Supplementary Table S3).

Under native ESCAT criteria, 40 of 57 assessed Level 1 biomarkers (70%) were concordant with the ESCAT tier assigned in the corresponding CPG, while 17 (30%) were discordant. After applying the predefined clinically aligned modifications, concordance increased to 49 of 57 biomarkers (86%) (Figure 1C),

The eight remaining discordant cases primarily reflected differences in the evidence considered, unclear documentation of the evidence base used by guideline committees, or apparent misalignment between the guideline-assigned tier and the type or strength of the available data (Supplemental Table S4). For example, NTRK2 fusions were assigned ESCAT Tier I-C in several ESMO CPGs despite being supported by only a single evaluable patient in basket trials, whereas our modified ESCAT assessment retained a Tier III-B classification due to insufficient gene-specific evidence [4].

### Inter-Reviewer Agreement and Variability

Perfect agreement among the three assigned reviewers were reached for 80 of 172 biomarkers (47%). In 12 of these cases, however, at least one reviewer requested additional discussion at a consensus meeting, indicating residual uncertainty despite initial concordance. Overall, 62% of final classifications were determined through consensus deliberation, either to resolve discrepant ratings or to address reviewer-initiated concerns.

Agreement between at least two of three reviewers was achieved for 143 biomarkers (83%), while 29 biomarkers (17%) had fully divergent initial ratings. Notably, 21 of these 29 biomarkers either mapped to ESCAT Tier X or did not map to any ESCAT tier under native criteria (Supplementary Table S2). Among the 37 biomarkers that did not map to any tier using strict criteria, reviewer agreement was particularly low (21%) underscoring the interpretive challenges in these cases and highlighting the limitations of the native ESCAT criteria to map certain evidence scenarios.

To quantify inter-rater reliability, we calculated Krippendorff’s alpha (α) using a nominal scale. The resulting α of 0.586 is consistent with tentative agreement and further illustrates the inherent variability in applying native ESCAT criteria to biomarker–therapy pairs.

## DISCUSSION

The ESCAT framework was developed to standardize the evaluation of biomarker-driven treatment decisions and support consistent, evidence-based application of precision oncology. In practice, however, ESCAT has not been systematically integrated into clinical sequencing workflows, and no openly accessible resource currently provides curated ESCAT tiers for routine clinical or research use. In this study, we mapped FDA-recognized OncoKB™ Level 1 biomarkers as of 29 September 2025, to ESCAT tiers using only rigorously curated evidence within OncoKB™ and identified specific evidence scenarios that are difficult to classify under native ESCAT criteria. Nearly one-quarter of Level 1 biomarkers could not be assigned to a native ESCAT tier, and reviewer agreement was modest, underscoring the interpretive challenges encountered by clinicians.

Our predefined, clinically aligned modifications resolved all previously unclassifiable biomarkers and increased concordance with ESCAT assignments in ESMO CPGs from 70% to 86%. The resulting openly accessible ESCAT-aligned set of standard care biomarkers provides a transparent, reproducible resource for molecular tumor boards, variant interpretation pipelines, and educational efforts aimed at harmonizing precision oncology practice. These empirical findings may inform future updates of the ESCAT framework and remain applicable even in settings that use complementary or alternative evidence-classification systems.

Prior disease-specific applications of ESCAT, particularly in breast and lung cancer, have demonstrated its clinical utility by enabling structured ranking of actionable biomarkers and the evidence supporting treatment decisions [5,6]. However, these efforts relied on expert panels performing comprehensive literature review and contextual judgement, underscoring the difficulty of deploying ESCAT systematically without curated resources or explicit guidance for ambiguous evidence scenarios.

### Inter-reviewer variability and implications for clinical practice

Inter-reviewer agreement when applying native ESCAT criteria was modest, with complete concordance for fewer than half of all independently reviewed biomarkers. Similar variability has been reported previously, where clinicians applying ESCAT to real-world evidence frequently assigned different tiers to the same biomarker–therapy pairs, even when using identical source material [7]. In our study, disagreement occurred predominantly in cases that ultimately proved unclassifiable under native ESCAT criteria, indicating that variability stemmed largely from structural constraints of the framework rather than subjective differences in interpretation. By isolating the evidence scenarios that generate the greatest ambiguity, our results highlight where additional methodological guidance or future ESCAT updates may have the greatest impact.

### Limitations of the current ESCAT framework

In our mapping, 22% of OncoKB™ Level 1 biomarkers could not be assigned to a native ESCAT tier. These unmapped cases reflected two recurrent evidence scenarios. First, single-arm trials reporting survival endpoints, often showing substantial clinical activity and supporting regulatory approval, cannot meet the MCBS requirements necessary for Tier I-B if quality-of-life data are absent [8]. Second, therapies demonstrating survival outcomes comparable, but not superior, to an alteration-matched standard care cannot be assigned Tier I-A, even when the biomarker’s actionability is well established [9]. Our predefined, clinically aligned modifications addressed these limitations and enabled consistent tier assignment in both scenarios.

### Alignment of ESCAT classifications with clinical practice standards

ESMO CPGs represent the most authoritative setting in which ESCAT tiers are formally applied by disease-specific expert committees. We therefore used CPG assessments as a practical benchmark for how ESCAT is used in real-world clinical decision-making. Our analysis shows that native ESCAT criteria frequently diverged from these real-world assignments, whereas the modified criteria substantially improved concordance.

The pattern of discordance is informative. First, most mismatches under native ESCAT occurred in the two evidence scenarios we identified as structurally not matching to an ESCAT tier, confirming that the challenges are intrinsic to the framework rather than due to inconsistent interpretation. Second, the few remaining discordant cases after applying the modified criteria primarily reflected differences in the evidence reviewed by guideline committees, occasional ambiguity regarding which studies informed the published ESCAT tier, or rare instances in which the assigned tier appeared inconsistent with the strength of cited data (Supplementary Table 4). Importantly, the increased concordance achieved with our modified criteria suggest that targeted, transparent methodologically adjustments can bring ESCAT more closely in line with how precision oncology is used in clinical practice.

### Biomarker granularity and composite evidence

Composite biomarkers, which group genes by shared biological processes such as homologous recombination repair, and variant-level distinctions pose additional challenges for ESCAT classification. Several OncoKB™ Level 1 genes supported by studies evaluating composite biomarkers did not reach high ESCAT tiers because gene- or variant-specific outcomes were unavailable or based on small numbers. This reflects a broader limitation of FDA-aligned OncoKB™ Level 1 designations, in which the clinical benefit observed in composite-marker trials is often driven by a small subset of well-established genes. Our conservative assignment approach is therefore warranted and aligns overinterpretation of limited subgroup data can lead to overly broad assumptions of treatment benefit for specific biomarkers [10].

### Strengths and limitations

A key strength of this work is the use of the rigorously curated, transparent evidence available in the OncoKB™, which ensured transparency, avoided selective literature inclusion and supported reproducibility. Independent review by multiple oncologists, combined with a structured consensus process, further strengthened the robustness of the classification.

Limitations include that reviewers were not always disease subspecialists, which may have limited nuance in trial interpretation, although random assignment reduced bias and prevented anchoring to practice patterns. Additionally, the reviewer panel was composed of clinicians from a single institution who regularly participated in molecular tumor board decision-making. Independent validation of these findings by external expert groups would further strengthen the generalizability of the results. By providing the mapped dataset via accessible OncoKB website (oncokb.org) we hope to facilitate independent evaluation and future multi-institutional evaluation.

Our analysis was confined to the evidence curated within OncoKB™, and while our modifications improved classification consistency, they represent pragmatic adaptations rather than formal revisions to the ESCAT framework.

## Conclusions

Under strict application of native ESCAT criteria which requires meeting the stringent criteria of the MCBS, a subset of clinically validated standard-of-care biomarkers remained unclassifiable because the supporting evidence did not fit any existing ESCAT tier. Introducing simple and transparent modifications to resolve previously unclassifiable cases we demonstrate that the modified framework improves alignment with ESMO CPGs. The resulting classifications, now available at oncoKB.org, constitute the first openly accessible resource for systematic ESCAT evaluation of standard care genomic biomarkers in precision oncology.

Beyond the operational modifications applied in this study, additional strategies may help address the observed mismatch between regulatory recognition of biomarker–drug associations and the current ESCAT evidence framework. These include explicit ESCAT guidance for biomarker–drug associations supported by registration-enabling single-arm trials, particularly in rare diseases or molecularly defined populations where randomized studies may not be feasible (such as Erdheim-Chester disease). A second potential area for clarification concerns therapies demonstrating outcomes comparable, but not superior to, an established standard of care precision oncology drug, which currently does not map to the existing ESCAT tiers despite demonstrated clinical utility.

Finally, future iterations of the framework may consider structured expert-committee adjudication for evidence scenarios that while clinically compelling, do not align with current ESCAT tiers. While these considerations extend beyond the scope of the present study, these analyses may help inform future iterations of the ESCAT framework

## Data Availability

All data produced in the present work are contained in the manuscript.

## ACKNOWLEGEMENTS

The authors would like to thank the entire OncoKB™ team for valuable scientific discussions and technical support.

